# A monitoring method to evaluate the accumulation of antimicrobial-resistance genes on plasmids harbored by antimicrobial-resistant bacteria distributed in environmental water

**DOI:** 10.1101/2025.06.16.25329729

**Authors:** Nobuyoshi Yagi, Sora Miyazato, Nguyen Quoc Anh, Bui Thi Mai Huong, Itaru Hirai

## Abstract

Antimicrobial-resistant bacteria (AMRB) distributed in the environment can cause human refractory infections. Frequently performed environmental metagenomics could not provide information on what types of AMRB accumulate what types of antimicrobial resistance genes (ARGs). Assuming to monitor AMRB in the environment, we selected 910 AMRB isolates using cefotaxime and ciprofloxacin from Vietnamese environmental water samples and subjected to 16S rRNA sequencing. It indicated that *Escherichia coli* (36.0%), *Citrobacter freundii* (21.4%), *Acinetobacter baumannii* (20.6%), and *Klebsiella pneumoniae* (19.7%) were dominant. Using *E. coli* as a model, we further analyzed AMRB isolates by phylogenetic analysis and whole-genome sequencing (WGS). The sequenced full-length *fimH* of the isolates were plotted with the already published *fimH* sequences on a phylogenetic tree. Considering phylogeny, 14 strains were subjected to WGS that indicated not only the number and type of ARGs but also the order of ARGs on the plasmid were confirmed in the analyzed *E. coli* isolates. More importantly, 3 of the 14 strains were *bla*_NDM-5_-positive, *i.e.*, carbapenem-resistant *E. coli*. These results suggest that our analytical procedure in this study is applicable as a monitoring method to understand in detail genetic characteristics of AMRB isolates in environmental water samples.

## Introduction

As in case that extended-spectrum β-lactamase (ESBL)-producing Enterobacteriaceae have been detected worldwide [1], it is supposed that certain types of AMRB are circulating among healthcare-associated facilities, communities, and the environment [2]. In many regions and countries, surveillance of AMRB is mainly conducted in healthcare-associated facilities. However, almost no region or country that maintains monitoring system for AMRB spreading in the environment, food or healthy individuals. This is likely due to a lack of analytical systems to monitor AMRB at a reasonable cost.

Regarding the AMRB spread in the environment, instead of isolating AMRB by conventional microbiological methods, ARGs have been detected in environmental water samples, such as river water or pond water. To detect ARGs in the environment, next-generation sequencing has been used in amplicon sequencing, environmental metabarcoding, metagenomics and so forth [3]. These techniques are powerful enough to detect most of the existing ARGs and determine the bacterial species contained in each sample tested. However, unless WGS of bacterial strains is performed, it is almost impossible to observe how many ARGs have been accumulated on plasmid(s) in the tested bacteria and how ARGs are arranged whether on plasmid or chromosome. Many studies have observed that various ARGs accumulated on plasmids that can be transferred beyond barriers among bacterial species. And it is regarded that the acquisition of plasmids carrying these multiple ARGs by antimicrobial-sensitive bacteria is one of the factors in the emergence of multidrug resistant bacteria (MRB) [4]. Therefore, it is essential to understand which and how many ARGs would be accumulated on plasmid and to analyze the genetic background of AMRB to study the distribution mechanism of AMRB.

It is possible that AMRB might circulate among healthcare-associated facilities, communities, the environment, etc. and function as a reservoir of ARGs. To support this, previous studies have reported that carbapenem-resistant and colistin-resistant bacteria, which are resistant to the last resort drugs, carbapenems and colistin, respectively, were detected in the community [5–9]. For these reasons, it is considered important to monitor AMRB in the environment as well as AMRB surveillance in healthcare-associated facilities.

Meanwhile, in our previous research, we established a WGS protocol, the Shallow-sequencing, for AMRB that could sufficiently reduce the cost per strain, and a way for presuming the molecular structure of plasmids harboring ARGs from the obtained genome sequence of AMRB [10]. The established protocol allows us to analyze a greater number of AMRB and to evaluate how and how many ARGs have been accumulated on the antimicrobial-resistance plasmids carried by AMRB. In this study, we applied the analytical protocol to consider what steps could be taken to observe AMRB spreading in the environments using environmental water samples collected from Vietnamese communities.

## Experimental Procedures

### Environmental water samples and isolation of AMRB

Environmental water samples were collected from rivers and ponds in the BaVi area, Hanoi city, Vietnam. Basically, 50 ml of environmental water was collected by 50 ml conical tube and bring back to the National Institute of Nutrition, Vietnam. Consequently, 12 environmental water samples were collected. One dip of each of the 12 collected environmental water samples was taken by the seed swab kits (Eiken Chemical Co., Ltd., Tokyo, Japan) and used for cultivation of MacConkey agar without any antibiotics. The grown bacteria were collected in 2 ml of trypticase soy broth (TSB) containing 10% glycerol and stored at −80 C°. One loop of the stocked bacteria was diluted with TSB from 10^-1^ to 10^-5^ and 100 μl of the diluted bacterial solution was inoculated on MacConkey agar containing 2 μg/ml of cefotaxime or 2 μg/ml of ciprofloxacin. The grown bacterial colonies, *i.e.*, AMRB strains, were collected in 96 well plates containing 150 μl of TSB containing 10% glycerol and stored at −20 C° until analysis.

### Amplification of full-length of 16S rRNA gene for identification of bacterial species

To identify bacterial species of the collected AMRB strains, full-length of 16S rRNA gene was amplified by gene specific primers (**Table 1**). Bacterial DNA was extracted by the conventional boiling method. Briefly, 1 ml of bacterial culture was centrifuged at 14,500 rpm for 1 min and the precipitated bacterial cells were resuspend in 0.5 ml of 10 mM Tris, HCl, pH8.0, 1mM EDTA buffer. After 5 min of incubation at 100 C°, the bacterial suspension was centrifuged at 14,500 rpm for 10 min. Consequent supernatant was used as templates for PCR after adjusting DNA concentration at 1.0 ng/μl. To amplify full-length of 16S rRNA, GoTaq Green Master Mix (Promega K.K., Tokyo, Japan) was used by following the manufacture’s instruction. Briefly, 1ng of the extracted bacterial DNA was used as the template in 10 μl of reaction mixture containing forward primer and one of the barcoded-reverse primers (**Table 1**). Thermal conditions for amplifying full-length 16S rRNA was as follows, an initial melt of 98 C° for 2 min, 30 cycles of 98 C° for 10 sec, 55 C° for 30 sec and 72 C° for 1min 45 sec, and a final extension at 72 C° for 7 min. Then the amplified DNA samples were purified by KAPA Pure Beads (Roche Sequencing Solutions, Inc., Pleasanton, CA) and subjected to the Nanopore sequencing (Oxford Nanopore Technologies, Oxford, UK).

**Table 1.**
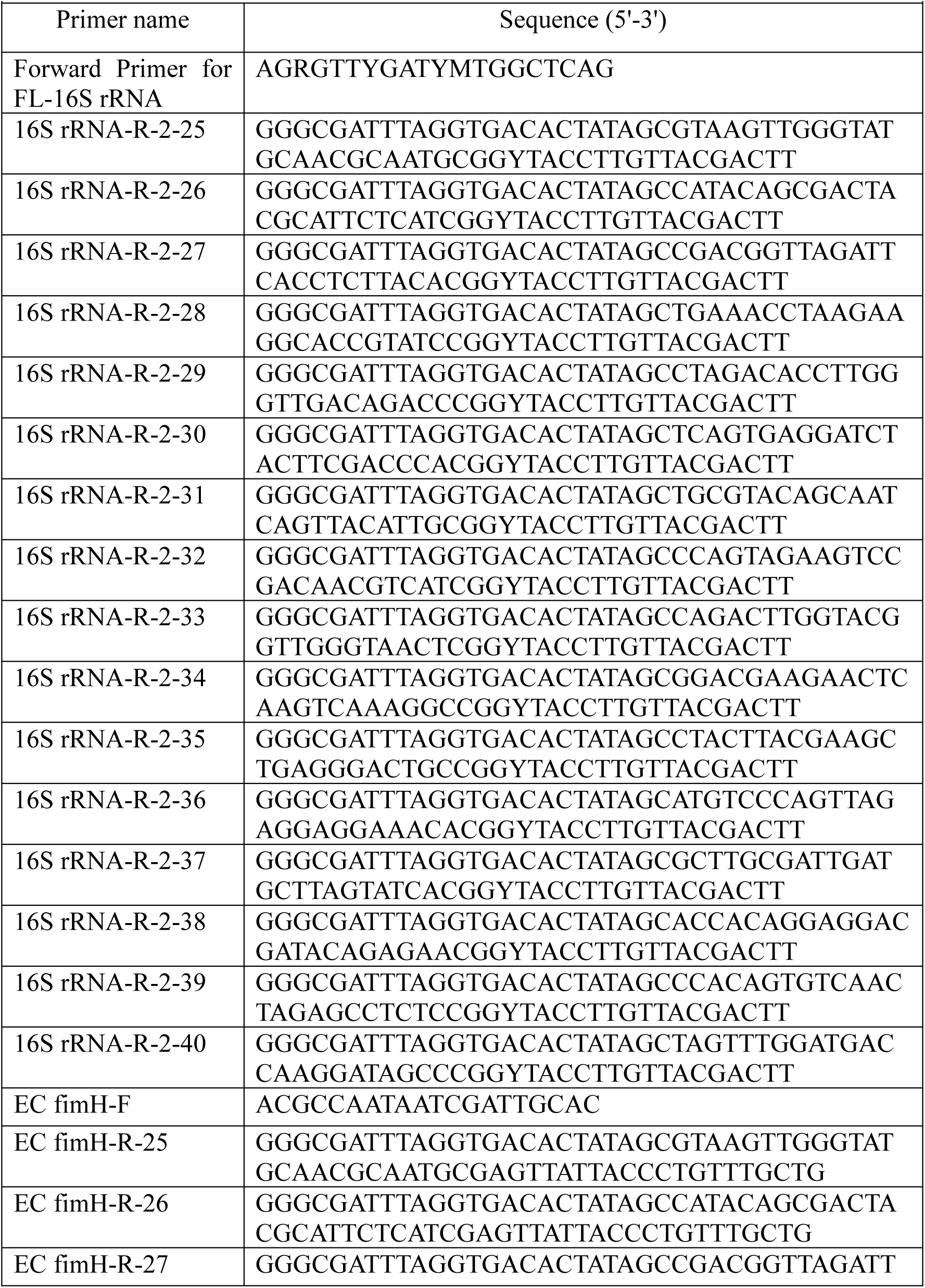
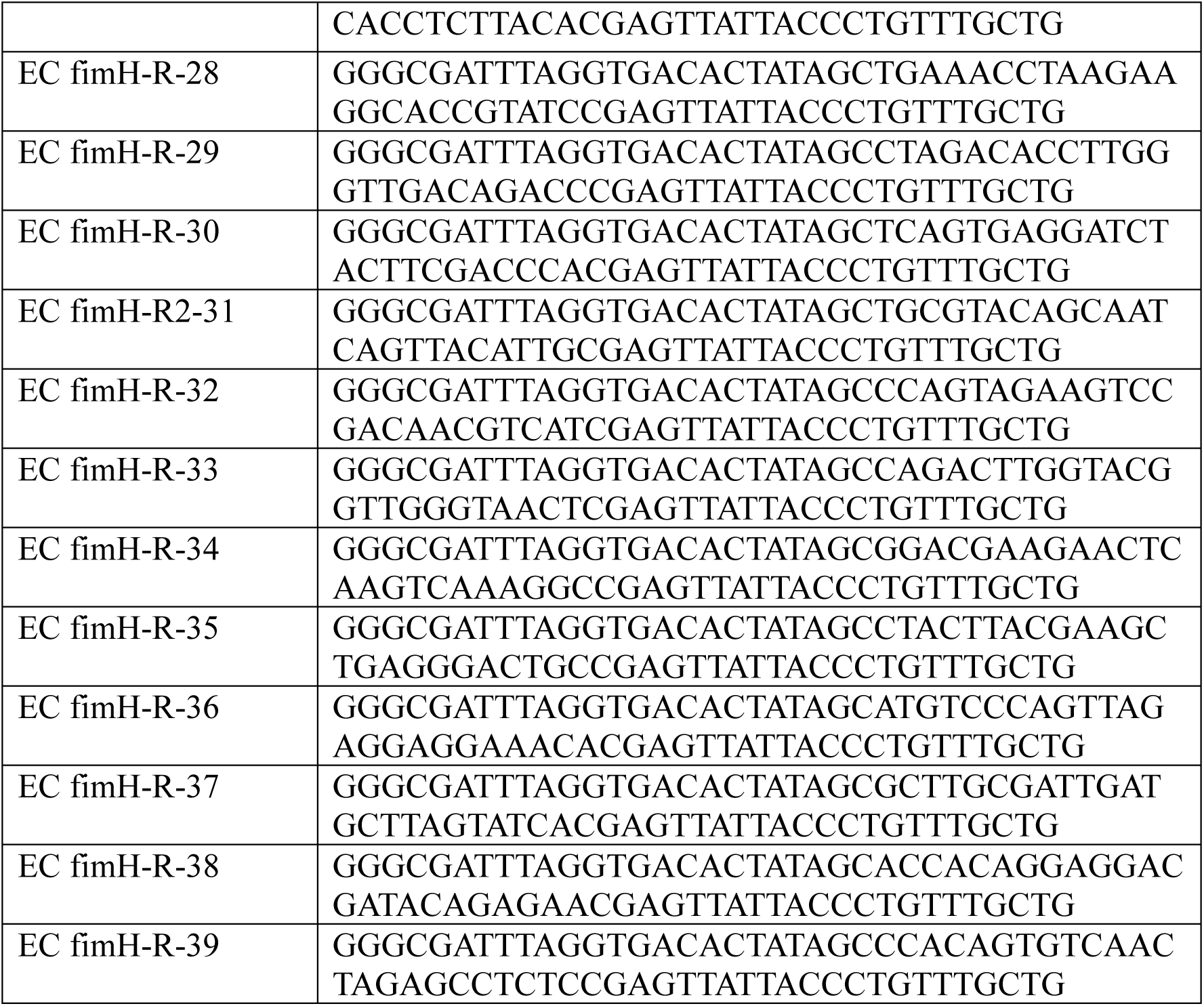
Sequences of primers in this study.

### Amplification of *E. coli* by *fimH* for phylogenetic classification

To amplify *fimH*, 1 ng of the extracted bacterial DNA was used as the template in 10 μl of GoTaq Green Master Mix reaction mixture containing forward primer and one of the barcoded-reverse primers (**Table 1**), and performed thermal cycles consist of an initial melt of 98 C° for 2 min, 30 cycles of 98 C° for 10 sec, 53 C° for 30 sec and 72 C° for 1min, and a final extension at 72 C° for 7 min. The amplified DNA samples were purified by KAPA Pure Beads by following the manufacture’s instruction and subjected to the Nanopore sequencing.

### Bacterial genomic DNA purification

Bacterial genomic DNA of the representative AMRB strains were purified by the Monarch Spin gDNA Extraction Kit (New England Biolabs., Ipswich, MA) by following the manufacture’s instruction.

### Library preparation and Nanopore sequencing

The purified amplified DNA or bacterial genomic DNA were subjected to Nanopore sequencing. For Nanopore sequencing 65 ng of the amplified DNA or 200 ng of the bacterial genomic DNA was used to prepare libraries by following the protocol of the Native Barcoding Kit 24 V14 provided by Oxford Nanopore Technologies. Nanopore sequencing was performed using the Flongle Flow Cell (R10.4.1).

### Sequence read cleanup

The obtained sequence reads were demultiplexed based on barcode sequences using Porechop [11] and the demultiplexed, *i.e.*, de-barcoded sequence reads were used for following analysis. The de-barcoded sequence reads were subjected to quality filtering with quality scores and read length. For 16S rRNA gene sequencing and WGS of AMRB strains, sequence reads with a quality score of at least 10 and a read length of 1.0 k bp or more were used. For *fimH* typing, only sequence reads with a quality score of at least 14 and a read length of 1.2 k bp or more were used in the analysis.

### Bacterial species identification using 16S rRNA sequences

Bacterial species identification based on the 16S rRNA gene sequence was performed using Emu [12]. Among the emu analysis results, only data with an “abundance” of 95% or higher and “estimated counts” of 100 or higher were selected.

#### *fimH* typing

The cleaned sequence reads were analyzed using Amplicon_Sorter [13] to obtain consensus *fimH* sequences of *E. coli* isolates obtained in this study. These consensus sequences were then analyzed with FimTyper [14] to determine the fimH type. All *fimH* sequences that were registered in the FimTyper database were clustered at a cut off value (99% identity). A phylogenetic tree was constructed using the clustered *fimH* sequences and the *fimH* sequences of the determined fimH types. MEGA11 was used for phylogenetic tree construction [15].

### WGS of AMRB isolates and data analysis

*De novo* assembly was performed using Flye [16] and the cleaned sequence reads originating from the representative AMRB strains. The obtained draft assemblies were polished using medaka (https://github.com/nanoporetech/medaka) to generate consensus sequences, *i.e.*, contigs. The generated contigs of the representative AMRB strains were subjected to identifying chromosome-derived open reading frames (ORFs) [17].

The ORFs of each representative AMRB strain were detected by blastn [18] and a ORF database included all ORFs that was constructed with PanTA [19] and 5,000 complete *E. coli* genome sequences retrieved from NCBI datasets. The detected and absent (not detected) ORFs were assigned a value of 1 and 0, respectively, collected in an ORF presence-absence table, and were converted to binary sequences for each *E. coli* genome sequence. The binary sequences were analyzed using the BINNGAMMA model in RAxML [20]. The resulting data were visualized using R and RStudio to construct an ORF tree.

ARGs and plasmid replicons were detected using AMRFinderPlus [21] and PlasmidFinder [22], respectively. The contigs, in which ARGs were detected, were assigned as plasmid-derived contigs using PLASMe [23].

Plasmid sequences were retrieved from the RefSeq database by keyword search using “complete sequence,” filtered by species: “bacteria”, genetic compartment: “plasmid”, and sequence length: 3,000 to 500,000 bp. BLAST searches were performed to identify plasmid sequences homologous to the detected plasmid-derived contigs. The plasmid sequence with the highest % identity and query cover to each plasmid-derived contig was designated as the reference plasmid sequence. The similarity among the plasmid sequences, the reference sequences, and the plasmid-derived contigs were assessed using BRIG [24]. In addition, the ARG cluster between the plasmid reference sequences and the plasmid-derived contigs was drawn using Clinker [25], based on GenBank files annotated with the BacAnt [26].

## Results

In this study, we isolated 910 bacterial colonies from the collected 12 environmental water samples and subjected to 16S rRNA amplification to identify bacterial species. Consequently, bacterial species of 791 (86.9%) of the 910 isolates were identified. *E. coli* and *K. pneumoniae* were detected both in the CTX-resistant and CIP-resistant isolates. Most of *C. freundii* and *A. baumannii* were mainly detected in CIP-resistant and CTX-resistant isolates, respectively (**Table 2**).

**Table 2.** Identified bacterial species of the isolates obtained from the ponds and rivers.

| Identified isolates | Total |  | CTX-resistant isolates |  |  | CIP-resistant isolates |  |  |
| --- | --- | --- | --- | --- | --- | --- | --- | --- |
|  | n | %<sup>1)</sup> | n | %<sup>1)</sup> |  | n | %<sup>1)</sup> |  |
| Identified isolates | 791 |  | 378 | 47.8 |  | 413 | 52.2 |  |
| Bacterial spp. | n | %<sup>1)</sup> | n | %<sup>2)</sup> | %<sup>3)</sup> | n | %<sup>2)</sup> | %<sup>3)</sup> |
| *Escherichia coli* | 285 | 36.0 | 169 | 59.3 | 44.7 | 116 | 40.7 | 28.1 |
| *Citrobacter freundii* | 169 | 21.4 | 10 | 5.9 | 2.6 | 159 | 94.1 | 38.5 |
| *Acinetobacter baumannii* | 163 | 20.6 | 162 | 99.4 | 42.9 | 1 | 0.6 | 0.2 |
| *Klebsiella pneumoniae* | 156 | 19.7 | 23 | 14.7 | 6.1 | 133 | 85.3 | 32.2 |
| *Acinetobacter. Pittii* | 3 | 0.4 | 2 | 66.7 | 0.5 | 1 | 33.3 | 0.2 |
| *Acinetobacter soli* | 8 | 1.0 | 8 | 100.0 | 2.1 | 0 | 0.0 | 0.0 |
| others | 7 | 0.9 | 4<sup>4)</sup> | 57.1 | 1.1 | 3<sup>5)</sup> | 42.9 | 0.7 |

<sup>1)</sup>, Ratios in the total number of the AMRBs.
<sup>2)</sup>, Ratios in the total number of the identified species.
<sup>3)</sup>, Ratios in the CTX- or CIP-resistant isolates.
<sup>4)</sup>, *Aeromonas caviae*
<sup>5)</sup>, *Enterobacter hormaechei*

When considering the operation of an AMRB environmental monitoring system, collecting detailed genetic characteristics of AMRB isolates is essential. In other words, WGS of AMRB is to be performed to understand what kind of AMRB is prevalent in the target area. However, various AMRB species are potentially detected in AMRB monitoring, and it is not always easy to determine which AMRB species and isolates should be the target of WGS. Practically, it is necessary to select AMRB species that can be related to damage to objects of interest such as humans or livestock. In this study, we selected *E. coli*, a bacterial species associated with human health damage as a model and analyzed using a feasible procedure in the AMRB monitoring system.

Firstly, we performed phylogenetic grouping by *fimH* of *E. coli* (**Fig. 1**) to assess which genetic lineages the *E. coli* strains obtained in this study were closest to lineages of previously reported strains. As shown in **Fig. 1** and **Supplementary Table S1**, a total of 285 *E. coli* isolates were classified into 8 *fimH* types. The numbers of strains contained in each *fimH* type, fimH24, fimH34, fimH40, fimH43, fimH54, fimH65, fimH167, and fimH520, were 86 (30.2%), 50 (17.5%), 14 (4.9%), 6 (2.1%), 25 (8.8%), 65 (22.8%), 8 (2.1%) and 1 (0.4%), respectively. For the remaining 32 (11.2%) *E. coli* isolates, the *fimH* type was not determined. We confirmed relative locations of the eight fimH types observed in this study on a phylogenetic tree constructed using sequence data obtained from the public database, Genbank (**Fig. 1**). It was found that these eight fimH types appeared to be positioned separately from each other on the phylogenetic tree. Therefore, in this study, we selected a total of 14 *E. coli* isolates by considering *fimH* type distribution and subjected to WGS.

**Fig. 1.**
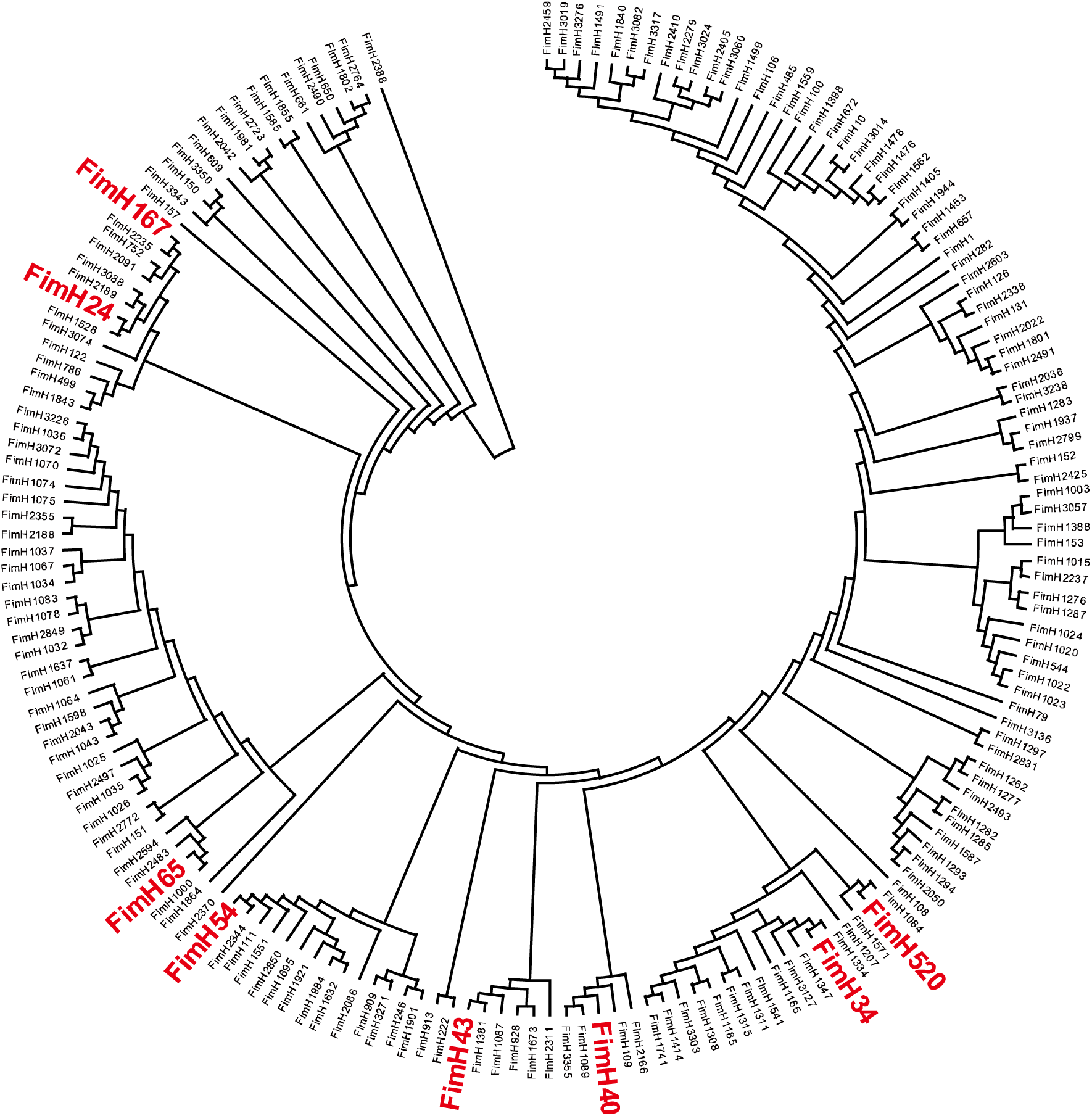
The *fimH* phylogenetic tree. The phylogenetic tree was constructed by the analyzed fimH sequence of the 285 *E. coli* isolates and representative *fimH* sequencies collected from the FimTyper database. The 185 *fimH* sequences were classified into 8 fimH types indicated by red color.

The genetic information of the 14 selected strains was collected by WGS. ORFs were extracted from the genetic information of the 14 analyzed strains. Binary sequences were obtained based on the presence or absence of ORF from the genetic information of the analyzed strains, and a phylogenetic tree was depicted combined with binary sequences obtained from the already published sequences with their phylogenetic groups (**Fig. 2** and **Supplementary Table S2**). Judging from the phylogenetic tree depicted by the ORFs, it seemed that the 14 selected *E. coli* isolates were classified into phylogenetic group A (9 isolates, 64.3%), B1 (2 isolates, 14.3%), and D (3 isolates, 21.4%).

**Fig. 2.**
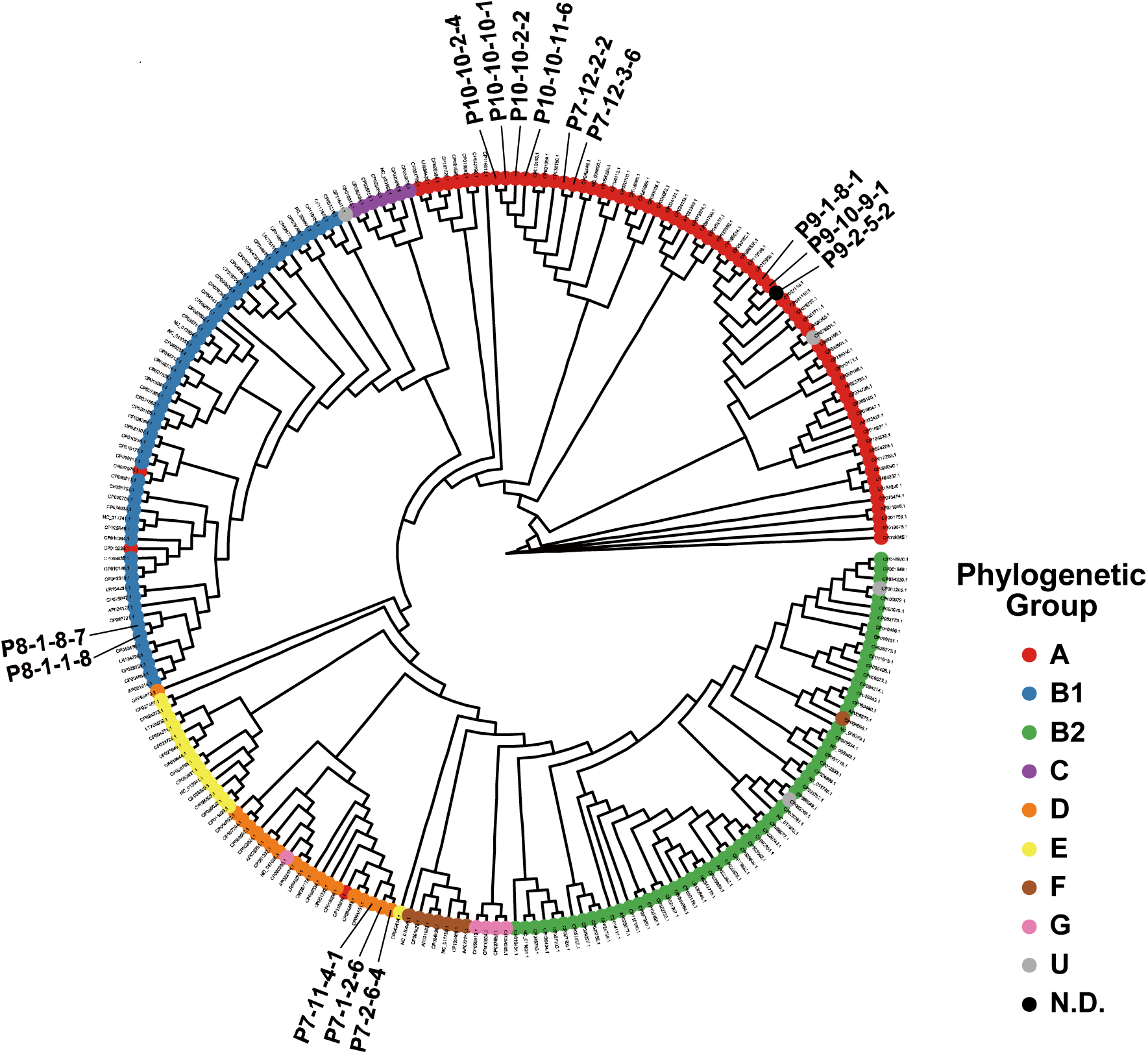
Comparison of genetic characteristics of *E. coli* isolates by an ORF tree. Genetic characteristics of the 14 analyzed *E. coli* isolates were compared with those of the 227 *E. coli* reference genome sequences collected from RefSeq by an ORF tree. For drawing ORF trees, an ORF database was constructed with PanTa and 5,000 complete *E. coli* genome sequences retrieved from NCBI datasets. The present and absent ORFs in each *E. coli* isolate genome was assigned a value of 1 and 0, respectively. Then it was converted to a binary sequence for each *E. coli* genome sequence. The binary sequences were analyzed using the BINNGAMMA model in RAxML and an ORF tree was drawn by R and RStudio.

In addition to the genetic lineages of the 14 *E. coli* isolates analyzed, the WGS provided information related to ARGs and plasmid replicons. Such as how many, and which and how ARGs could be accumulated, and which plasmid(s) were carried by the analyzed *E. coli* isolates (**Table 3**). In this study, any constructed contig that contained at least one ARG, *i.e.*, AMR Contig, was subjected to BLAST search to clarify whether the contig belonged to chromosome or plasmid. As shown in **Table 3**, 34 contigs were constructed from the 14 *E. coli* isolates sequenced. BLAST search indicated 8 (23.5%) and 26 (76.5%) of the 34 contigs were classified into chromosome and plasmid, respectively. It was suggested that each of the analyzed *E. coli* isolates possessed one to three plasmids, and that each plasmid carried 2 to 13 ARGs. Three *E. coli* isolates (P7-1-2-6, P7-2-6-4, P7-11-4-1) possessed *bla*_CTX-M_s and *bla*_DHA-1_, in addition to *bla*_NDM-5_ (**Table 3**, **Fig. 3**). The WGS indicated gene clusters in which the detected ARGs were arranged on the plasmids in the order shown in **Table 3**.

**Fig. 3.**
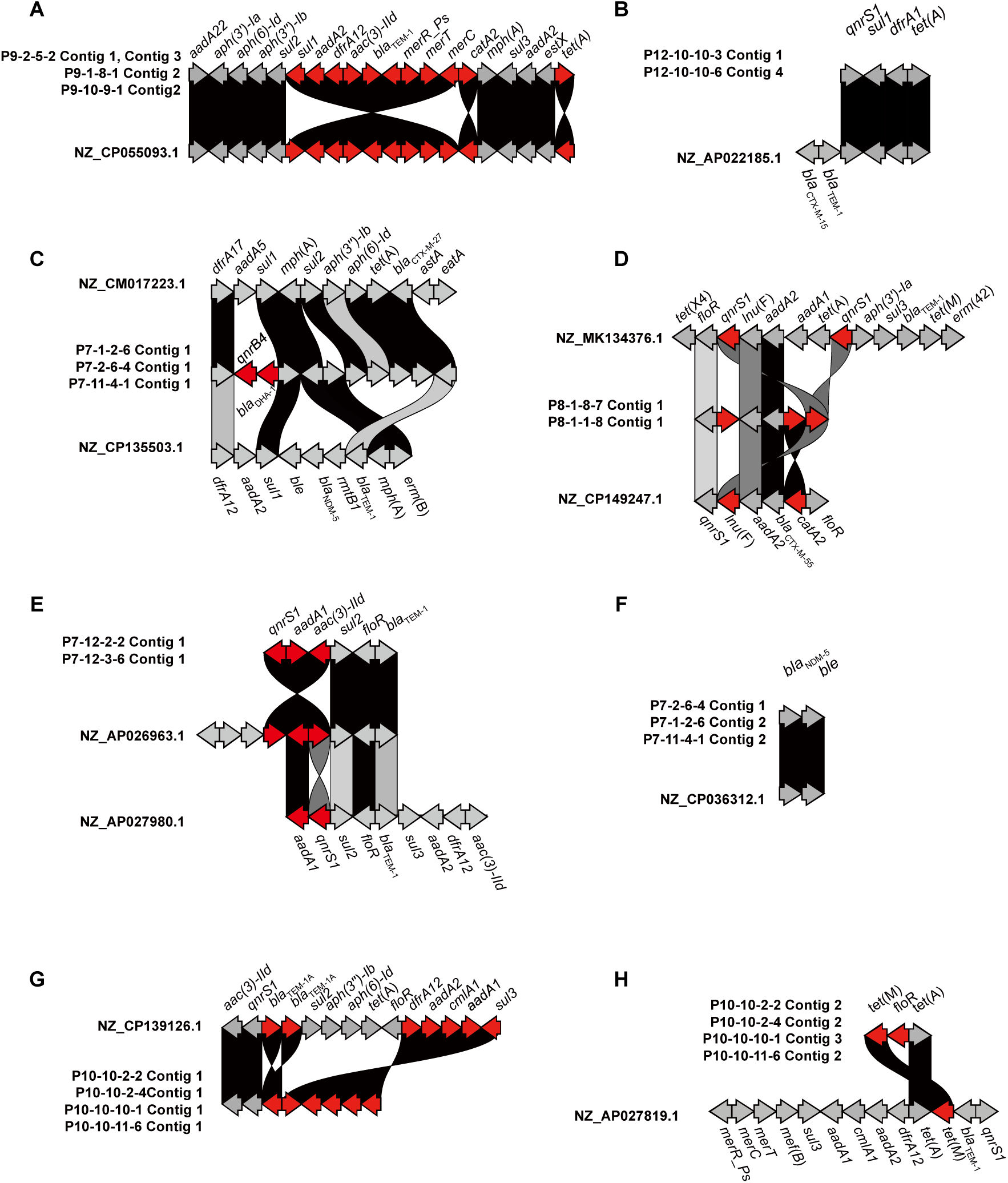
Comparison of gene clusters detected in the whole genome information of the 14 analyzed *E. coli* strains. The gene cluster plots were drawn by clinker to compare the order of ARGs, to simplify comparison of the ARGs arrangement, the gene scale was not considered. Red arrow means the difference between plasmid-derived contigs and reference sequences. **A**, P9-2-5-2 Contig 1and Contig 3, P9-1-8-1 Contig 2, P9-10-9-1 Contig2, **B**, P12-10-10-3 Contig 1, P12-10-10-6 Contig 4, **C**, P7-1-2-6 Contig 1, P7-2-6-4 Contig 1, P7-11-4-1 Contig 1, **D**, P8-1-8-7 Contig 1, P8-1-1-8 Contig 1, **E**, P7-12-2-2 Contig 1, P7-12-3-6 Contig 1, **F**, P7-2-6-4 Contig 1, P7-1-2-6 Contig 2, P7-11-4-1 Contig 2, **G**, P10-10-2-2 Contig 1, P10-10-2-4Contig 1, P10-10-10-1 Contig 1, P10-10-11-6 Contig 1, **H**, P10-10-2-2 Contig 2, P10-10-2-4 Contig 2, P10-10-10-1 Contig 3, P10-10-11-6 Contig 2.

**Table 3.**
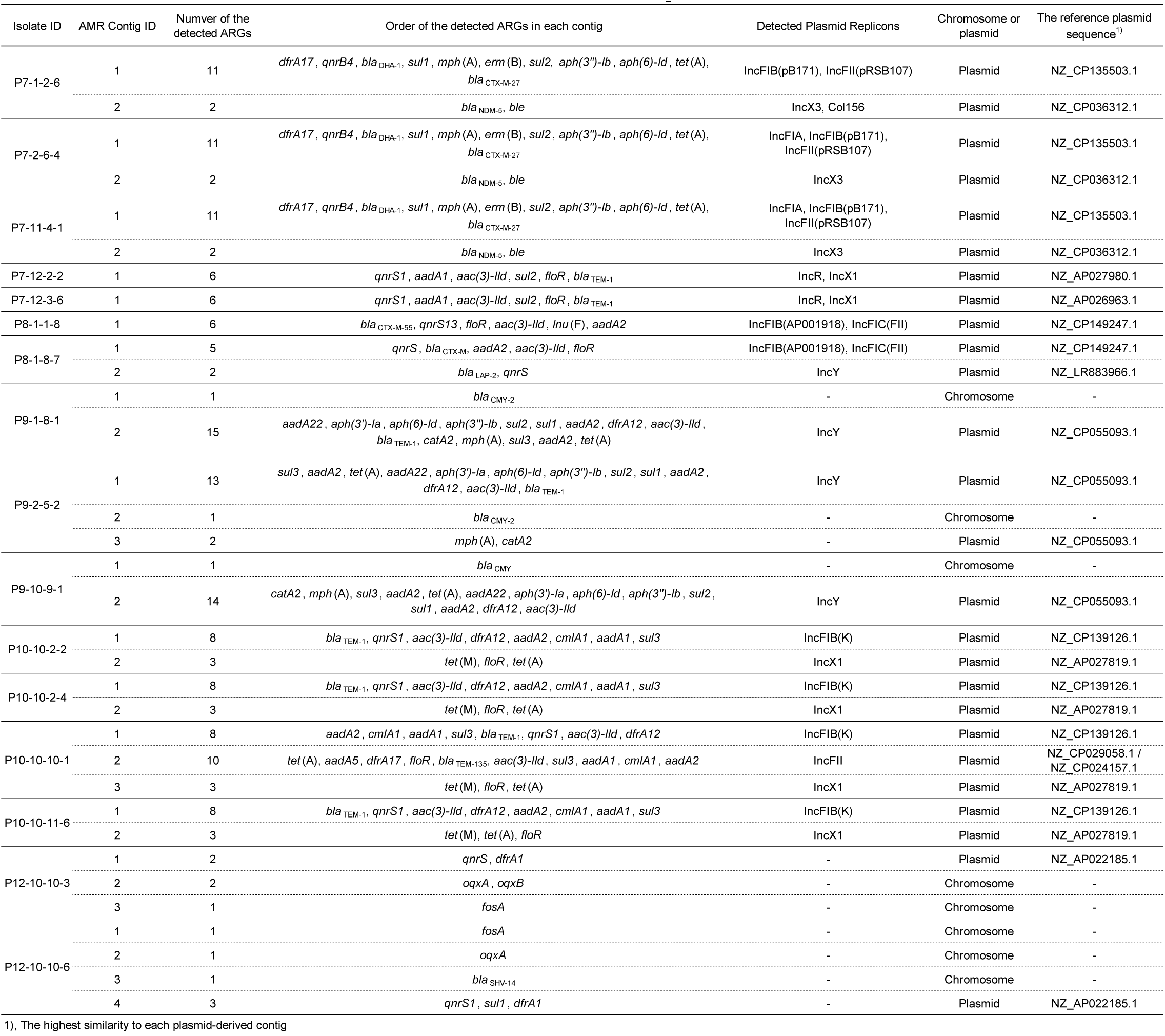
Detected ARGs and their order on the constructed contigs.

To understand plasmid structures, homologous plasmid sequences to each plasmid-derived contigs of the representative *E. coli* isolates were identified by BLAST search among the 22,089 plasmid sequences collected from the RefSeq database. The he highest similar plasmid sequence to each plasmid-derived contig was designated as the reference plasmid sequence (**Table 3**). By the BLAST search using plasmid-derived contigs as a query, it was revealed that the query cover rates of the plasmid-derived contigs against the reference plasmid sequences were between 74% to 100%. On the other hand, the coverage of reference plasmid sequences by each plasmid-derived contig was different by the reference plasmid sequences ranging from 34% to 100% (**Fig. 4**). Especially, plasmids carrying *bla*_NDM-5_ exhibited particularly high similarity, with over 200 sequences retrieved from the RefSeq database showing 100% query coverage rate, implying highly conserved plasmid structures.

**Fig. 4.**
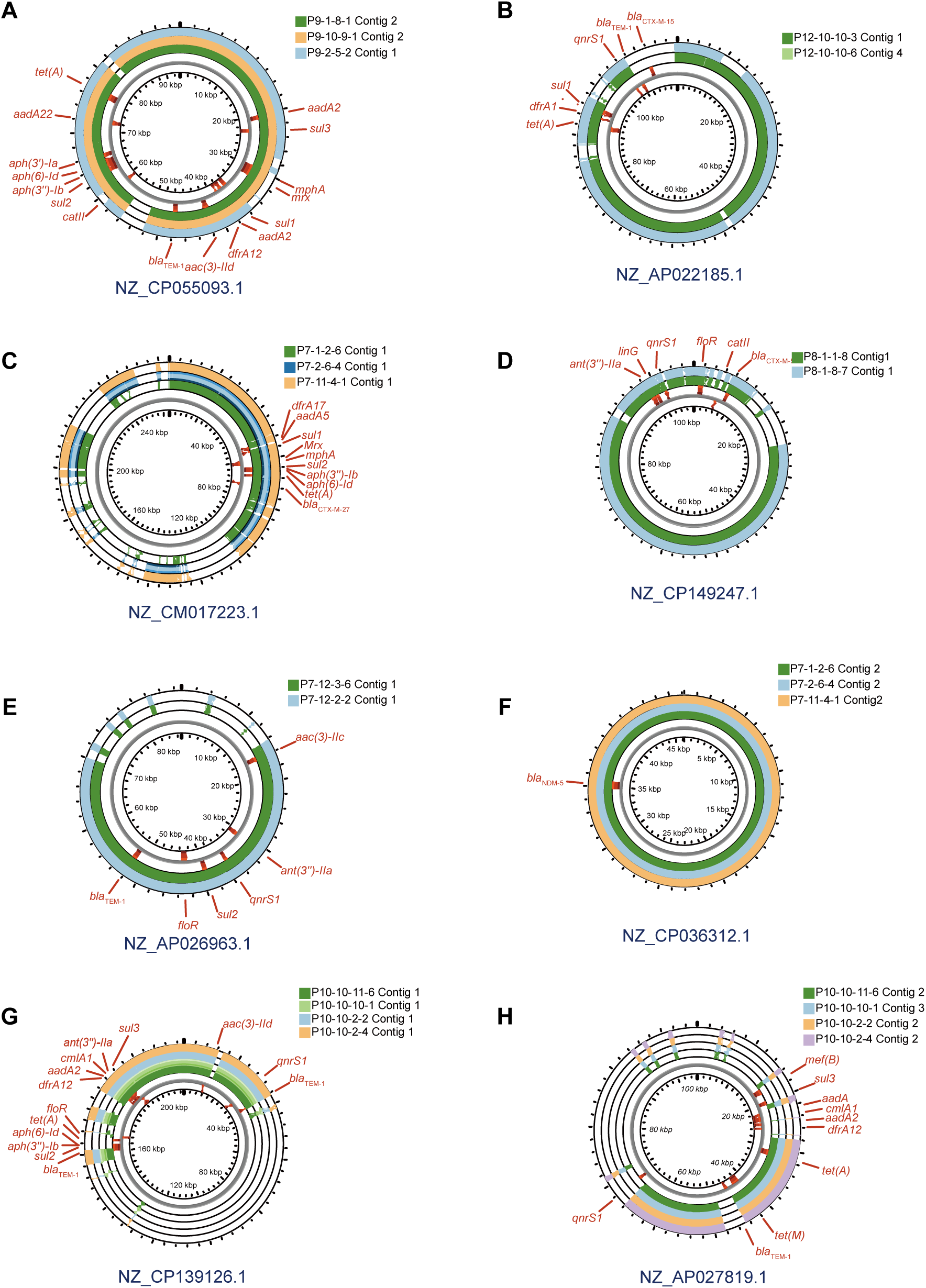
Comparison of plasmid sequences. Sample sequences were compared with homologous plasmid sequences detected from the plasmid databases. These circular maps were drawn by the Proksee website. ARGs locations were drawn by CARD in Proksee.

As mentioned above, WGS and subsequent data analysis not only confirmed the genetic background of the bacterial strains but also suggested that plasmids carrying different ARGs and replicons may have different plasmid evolution.

## Discussion

In antimicrobial-resistance surveillance conducted by healthcare-associated facilities, the target AMRB and ARGs have been generally selected by considering whether bacterial species could cause health hazards to humans. Regarding the environment, AMRB surveillance or monitoring has been almost completely neglected in many countries and regions for a long time. As a result, it has not been well explained what types of AMRB and ARGs are distributed in the environment. Currently, environmental metagenomics using next generation sequencer is enabling us to clarify the distribution of ARGs in the environment [27, 28]. However, environmental metagenomics have not been able to answer questions such as which bacterial species had become AMRB or how many ARGs had been accumulated in each antimicrobial-resistant bacterium.

This study proposes how to monitor AMRB distributed in environmental water. Our proposed monitoring method consisted of four parts. Namely, 1) selection of AMRB strains by solid media containing antibiotics, 2) bacterial species identification, 3) phylogenetic classification of AMRB strained for further analysis, and 4) WGS of selected AMRB strains and analysis of data obtained from WGS. In this study, we investigated how this proposed procedure was used to monitor AMRB in the environmental water by using *E. coli* strains as a model. Although the presence of Gram-positive bacteria in environmental water cannot be denied, it is important to observe the dynamics of Gram-negative bacteria when considering their impact on human health. Therefore, this study focused on Gram-negative bacteria.

Firstly, two antibiotics, CTX and CIP, were used in this study to select drug-resistant bacteria present in environmental water. CTX is designated by the CLSI as an antibiotic for screening ESBL-producing bacteria [29]. In this study, in addition to ESBL-producing bacteria, carbapenem-resistant bacteria, *i.e.*, *bla*_NDM-5_-positive *E. coli* strains, were also selected in CTX-containing media. Resistance to CTX occurs when bacteria acquire *bla*_CTX-M_ [30], carbapenemase producing genes including *bla*_KPC_, *bla*_OXA-48_, *bla*_IMP,_ and *bla*_NDM_, and drug efflux pump genes [7]. CTX-M type β-lactamase producing genes, *bla*_CTX-M_ is classified into several groups based on differences in its sequence, and it is known that there are regional differences in the *bla*_CTX-M_ groups possessed by ESBL-producing bacteria. Similarly, there are regional differences in carbapenemase producing genes, for example, *bla*_NDM_ is common in South Asia and Southeast Asia, and *bla*_IMP_ is commonly detected in Japan [7]. Our data suggested that it is possible to detect not only ESBL-producing bacteria but also various types of carbapenem-resistant bacteria by using CTX-containing media.

CIP was also used to select AMRB that are distributing in the environmental water samples. In general, bacteria become resistant to new quinolones through mutations in DNA gyrase genes or topoisomerase genes, or through the acquisition of quinolone resistance genes such as *qnrA*, *qnrB*, and *qnrS* [31, 32]. The proportion of bacteria resistant to new quinolones is gradually increasing, and AMRB isolated from the environment are often resistant to new quinolones as well. Furthermore, bacteria resistant to new quinolones often also carry other ARGs.

At a stage where AMRB monitoring has not yet been performed, it is almost impossible to determine which AMRB species should be monitored. Possibly, both Gram-positive and -negative bacteria would exist as AMRB in the environment. In this study, MacConkey agar medium was used to monitor Gram-negative bacteria. It should be decided whether Gram-positive or -negative AMRB were to be monitored depending on the purpose, *e.g.*, human health or livestock farming, and the area being monitored. In any case, when monitoring, the spread of AMRB species contained in the collected sample could be confirmed by analyzing the grown strains without deciding on the species to be monitored.

To identify the bacterial species, full-length 16S rRNA was determined by amplicon sequencing using Nanopore sequencer [33]. In this study, we hired 16 kinds of barcoded primers to amplify full-length 16S rRNA for Nanopore sequencing. By our results, the full-length analysis of 16S rRNA using one Flongle cell enabled simultaneous identification of at least 400 bacterial strains. As shown in **Table 2**, various AMRB species were detected, suggesting this study’s species identification step was effectively done.

It is essential to understand the genetic characteristics of AMRB species detected by AMRB monitoring through WGS. Because AMRB monitoring requires a lot of labor and expense, WGS must be performed as efficiently as possible. The genetic diversity of AMRB that is spreading in the environment is not well investigated, and there is few information on how to select target AMRB strains for WGS. In this study, phylogenetic grouping was performed to select strains for WGS. Phylogenetic grouping methods are well established for some species, such as *E. coli*, but for many species, it has not yet been enough. For instance, not all the genes of the conventional multilocus sequence typing (MLST) scheme are not always amplified in certain AMRB strains especially obtained from the environments [34, 35], and consequently sequence types of these AMRB strains would not be determined. And in many bacterial species, gene(s) that can be used to easily classify genetic lineages, such as the *fimH* in *E. coli*, has not been found. Therefore, for bacterial species other than major species such as *E. coli*, it is necessary to establish a method that can perform phylogenetic classification like *E. coli*.

The WGS performed in this study revealed many points, such as the relative position of the AMRB strain on the phylogenetic tree (**Fig. 2**), the location of carried ARGs, *i.e.*, on chromosome or on plasmid, and line up of ARGs, and the extent to which the AMRB strain had accumulated ARGs (**Table 3**).

Regarding plasmids carrying ARGs, as the results of this study show, it was possible that the evolution of plasmids differ depending on ARGs and plasmid replicons detected in the plasmids. To understand the evolution of plasmid carrying ARGs, it is important to observe what kinds of ARGs were carried by plasmids, and how genetic structure of plasmids isolated from many AMRB strains change. Then, comparison of genetic characteristics of plasmids carrying ARGs to others might make it possible to discuss how ARGs accumulate on the plasmids. In the environmental monitoring of AMRB, by creating a database including sample collection time and geographical information and observing the evolution of plasmids, it is highly possible to consider the temporal and geographical distribution of ARG-carrying plasmids. These things cannot be achieved with environmental metagenomics, which are currently commonly used.

As mentioned above, this study proposed a method for an AMRB monitoring system for environmental water. Regarding the cost of AMR monitoring that we proposed in this study, the Flongle cell (Oxford Nanopore Technologies, FLO-FLG114) used in this study cost US$ 918.00 for 12 cells, namely, each cell costs US$ 76.5. In addition, for 16S rRNA sequencing and *fimH* sequencing, more than 400 samples could be simultaneously analyzed with one Flongle cell, indicating it costs US$ 0.19 per sample. As for WGS, since 8 to 10 AMRB strains can be analyzed per one Flongle cell, it costs approximately US$ 7.65 per each AMRB strain. If the MinION cell that costs US$ 800 (Oxford Nanopore Technologies, FLO-MIN114) is used for the analysis, at most 48 Gb, that equivalent to more than 20 Flongle cells, could be analyzed when using MinION Mk1D. Therefore, it is suggested that the cost of AMR monitoring proposed in this study can be relatively low and not be a heavy burden for maintaining AMR monitoring.

However, there are some points to consider when installing the AMRB monitoring system. The first is how to determine the monitoring area. In many cases, there could be many factors to consider when setting up the monitoring area, and these would vary depending on the area. The second is how to determine how frequently and how many cases (samples) to monitor. This is also difficult to determine when the spread of AMRB is unknown in most areas of environmental water. In any case, it is important to conduct as much AMRB monitoring as possible and collect basic information using a relatively inexpensive method such as the one proposed in this study that could collect genetic information on AMRB.

## Data Availability

All data produced in the present study are available upon reasonable request to the authors.

## Acknowledgments

This work was funded by the KAKENHI 23K09672 to IT and 24K20197 to NY.

## Data availability statement

The whole genome sequencing data are openly available under NCBI BioProject number PRJDB20146.

## Author contributions

**Nobuyoshi Yagi**: conceptualization, data curation, formal analysis, funding acquisition, investigation, methodology, writing – original draft preparation. **Sora Miyazato**: formal analysis, investigation. **Nguyen Quoc Anh**: resources, writing – review & editing. **Bui Thi May Huong**: project administration, resources, writing – review & editing. **Itaru Hirai**: conceptualization, data curation, formal analysis, funding acquisition, investigation, methodology, project administration, writing – original draft preparation, writing – review & editing.

